# Evaluation of ELISA tests for the qualitative determination of IgG, IgM and IgA to SARS-CoV-2

**DOI:** 10.1101/2020.05.24.20111682

**Authors:** Colavita Francesca, Brogi Alessandra, Lapa Daniele, Bordi Licia, Matusali Giulia, Meschi Silvia, Marsella Patrizia, Tesi Giulia, Bandini Tommaso, Di Caro Antonino, Capobianchi Maria Rosaria, Castilletti Concetta, on behalf of the INMI Covid-19 laboratory team

## Abstract

Serological assays for anti-SARS-CoV-2 antibodies are now of critical importance to support diagnosis, guide epidemiological intervention, and understand immune response to natural infection and vaccine administration. We developed and validated new anti-SARS-CoV-2 IgG, IgM and IgA ELISA tests (ENZY-WELL SARS-CoV-2 ELISA, DIESSE Diagnostica Senese S.p.a.) based on whole-virus antigens. We used a total of 553 serum samples including samples from COVID-19 suspected and confirmed cases, healthy donors, and patients positive for other infections or autoimmune conditions. Overall, the assays showed good concordance with the indirect immunofluorescence reference test in terms of sensitivity and specificity. Especially for IgG and IgA, we observed high sensitivity (92.5 and 93.6%, respectively); specificity was high (>96%) for all antibody types ELISAs. In addition, sensitivity was linked to the days from symptoms onset (DSO) due to the seroconversion window, and for ENZY-WELL SARS-CoV-2 IgG and IgA ELISAs resulted 100% in those samples collected after 10 and 12 DSO, respectively. The results showed that ENZY-WELL SARS-CoV-2 ELISAs may represent a valid option for both diagnostic and epidemiological purposes, covering all different antibody types developed in SARS-CoV-2 immune response.

## Introduction

The frontline diagnostic response to the SARS-CoV-2 pandemic is represented by RT-PCR test performed on nasopharyngeal swabs or other upper respiratory tract specimens (**1**). In this phase of COVID-19 (coronavirus disease 2019) pandemic, there is a growing interest in antibody testing to determine the immune status against SARS-CoV-2 in convalescent and suspected cases as well as in the general population. In fact, serological investigation is crucial to better understand the host response to the virus, support diagnosis and help provide epidemiological estimates of virus spread (**2, 3**). Several serological assays are now becoming available and dozens of tests, including rapid tests, with variable degree of sensitivity and specificity are already on the market (**4**). To identify reliable laboratory diagnostics for SARS-CoV-2, validation on patient samples is necessary. Most of the available tests detect specific IgM and IgG, while IgA response is rarely considered, although representing the mucosal immunity (i.e. respiratory compartments) protecting the main sites of entry and replication for SARS-CoV-2 (**1**).

Here, we report the results of a study aimed at establishing the performances of a set of new commercial immunoenzymatic assays (ELISA) detecting specific anti-SARS-CoV-2 IgM, IgG and IgA antibodies. The work was carried out in the framework of a scientific collaboration established between DIESSE Diagnostica Senese S.p.A. (Siena) and the National Institute for Infectious Diseases (INMI) “L. Spallanzani” IRCCS.

## Methods

The ENZY-WELL SARS-CoV-2 IgM, IgG and IgA tests have been developed by DIESSE Diagnostica Senese S.p.A. and CE marked. These are based on the native antigen obtained from Vero E6 cells infected with the SARS-CoV-2, strain “2019-nCoV/Italy-INMI1” (EVAg Ref-SKU: 008V-03893).

A total of 553 anonymized serum samples were used to assess the performance of the ENZY-WELL tests. Of these, 115 were residual serum samples from 104 suspected and confirmed cases for SARS-CoV-2 infection, collected between February and April 2020, and previously evaluated with the reference in-house test established at the Laboratory of Virology at INMI, based on indirect immunofluorescence assay (IFA) detecting IgG, IgM and IgA. Fifty of these samples were patients with confirmed diagnosis by molecular testing. The anti-SARS-Cov-2 IFA results were as follows: IgA (64/110 positive), IgM (69/115 positive) and IgG (71/115 positive). Three hundred fifty three serum samples were retrieved from a retrospective collection of samples from healthy individuals collected in the pre-pandemic time (2016, 2017, and January 2019) and assumed to be SARS-CoV-2 negative. A group of 21 samples from patients who had been diagnosed with seasonal human coronaviruses (HKU, NL63, 229E), and 64 found positive for other infections (i.e. Mycoplasma, Cytomegalovirus, Adenovirus, Respiratory Syncitial virus, Influenza A, Influenza B, SARS-CoV) or interfering factors (i.e. as anti-nuclear antibodies and rheumatoid factor) was included in the analysis, to estimate cross-reactivity.

ELISA tests were performed on 1:100 serum duplicate dilution. For IgM ELISA, samples were treated with sorbent for IgG depletion. Serum dilutions were added to microwells coated with the whole-virus crude extract and incubated for 1 hour at room temperature. Then, microwells were washed and and anti-human IgG, IgM and IgA monoclonal antibodies, labeled with peroxidase, in phosphate buffer containing phenol 0.05% and Bronidox 0.02%, were added to the microwells (100 μL/well) and incubated at room temperature for 1 hour. After a second washing step, TMB substrate (100 μL/well) incubation was performed for 15 minutes, followed by addition of stop solution (100 μL/well). Optical density (OD) was read within 30 minutes at 450 nm with 620 nm subtraction, using Synergy™ HTX Multi-Mode Microplate Reader (BioTek Instruments, USA).

Internal quality controls were included in each run: one positive control containing anti-SARS-CoV-2 antibodies; one negative control not containing anti-SARS-CoV-2 antibodies; a cut-off (CO) control with a known concentration of anti-SARS-CoV-2 antibodies. The latter is used to calculate the Index value (ratio between the OD of the target sample and the CO) of the samples under evaluation. A blank control was also included in each run and values <0.150 contribute in confirming the quality of the test and procedures. Samples were considered positive when the Index is ≥1.0.

IFA, used as reference test, was performed using slides prepared in-house with Vero E6 cells infected with SARS-CoV-2 isolate (2019-nCoV/Italy-INMI1), as described elsewhere (**5**). Serum samples were tested for specific IgM, IgA and IgG using 1:20 screening dilution. FITC-conjugated anti-human IgM, IgA and IgG antibodies (Euroimmun, Germany) were used as secondary antibody and Evans Blue as cell counterstain. Antibody titres were obtained with titration by limiting dilution and expressed as the highest serum dilution presenting fluorescent antibody staining.

The diagnostic performance of ENZY-WELL SARS-CoV-2 ELISAs was calculated measuring sensitivity and specificity on the panel of serum samples above described. Concordance, k coefficient and Spearman correlation antibody levels amongst IFA and ENZY-WELL assays were calculated with respect to IFA. The statistical analysis was performed using GraphPad Prism version 7.00 (GraphPad Software, La Jolla California).

The study was approved from the INMI Ethical Board. Informed consent was not required as human samples used for this work were residual samples from routine diagnostic activities, patients’ data remained anonymous, and the results obtained with the new tests were not used for the clinical management of the patient.

## Results

On the samples from suspect and confirmed SARS-CoV-2 infection cases, ENZY-WELL SARS-CoV-2 IgG and IgA kits showed a good sensitivity (92.5 and 93.6%, respectively) and specificity (91.7% and 97.9%, respectively) as compared to the reference IFA test; on the contrary, IgM test showed lower sensitivity and specificity (87.7% and 88.0%, respectively, **Table 1**). On the whole, the agreement with IFA was excellent for IgG and IgA ENZY-WELL tests (92.7% and 95.4% concordance, respectively; k coefficient =0.84 and 0.9), while the agreement for IgM was lower, although still good (87.8% concordance and k coefficient =0.76).

**Table 1:** Diagnostic performance of ENZY-WELL SARS-CoV-2 IgG, IgM and IgA ELISAs as compared to IFA established on samples from suspect COVID-19 cases.

|  | ENZY-WELL<br>SARS-CoV-2 IgG | ENZY-WELL<br>SARS-CoV-2 IgM | ENZY-WELL SARS-CoV-2 IgA |
| --- | --- | --- | --- |
| **Sensitivity** | 92.5%<br>(CI 95%, 83.7 – 96.8) | 87.7%<br>(CI 95%, 77,5 - 93,6) | 93.6%<br>(CI 95%, 84,8 - 97,5) |
| **Specificity** | 91.7%<br>(CI 95%, 80.4-96.6) | 88.0%<br>(CI 95%, 76.2-94.4) | 97.9%<br>(CI 95%, 88.9-99.6) |
| **Concordance** | 92.7% | 87.8% | 95.4% |
| **k coefficient** | 0.84 | 0.76 | 0.9 |

The graphs in Figure 1 show the significant correlation of antibody levels amongst IFA and ENZY-WELL assays, as well as concordant and discordant results in the four quadrants (Figure 1).

**Figure 1.**
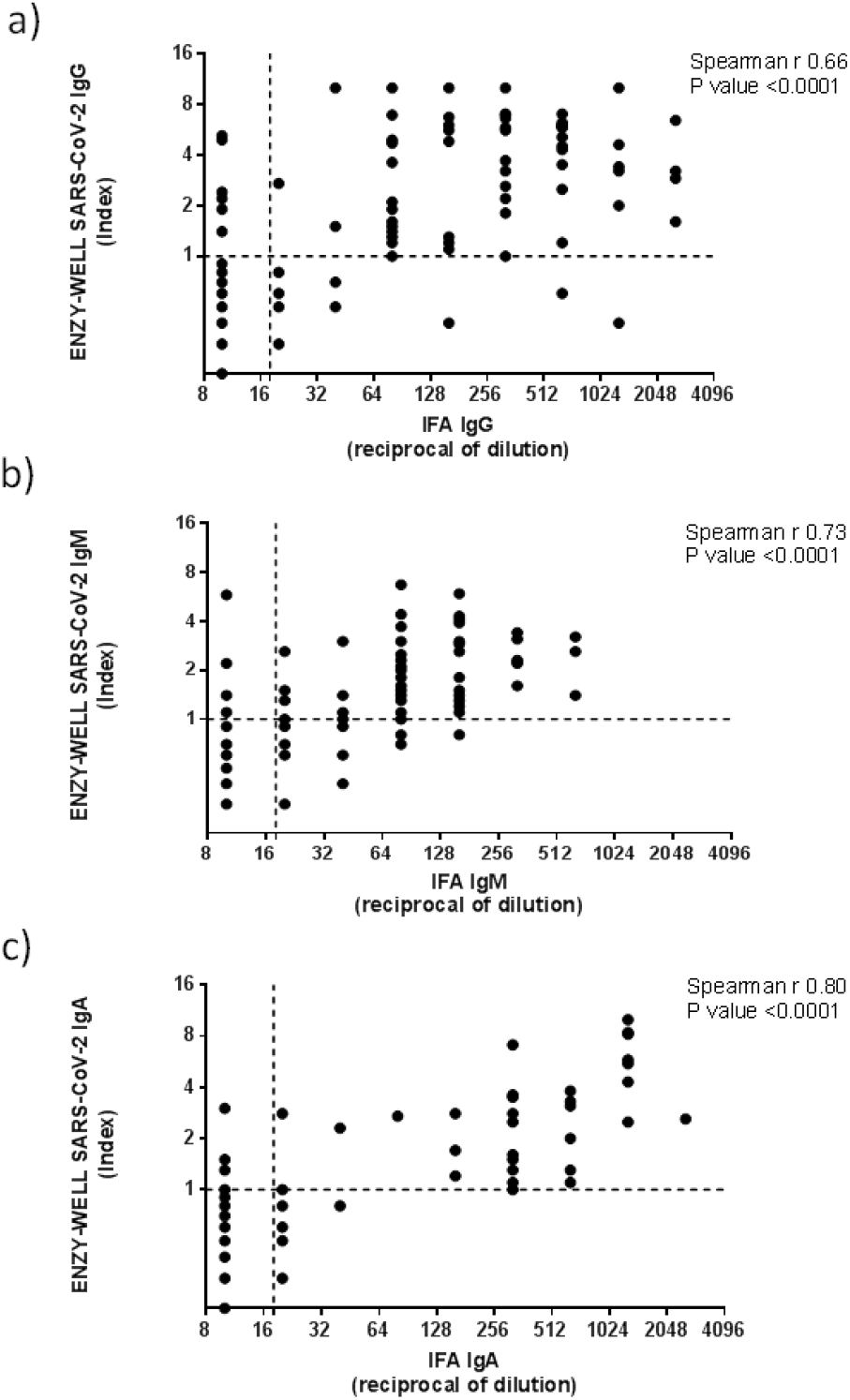
Correlation of antibody levels amongst IFA and ENZY-WELL assays. Antibody titres obtained from IFA are expressed as the highest serum dilution presenting fluorescence; levels obtained from ENZY-WELL ELISAs are expressed as Index value (ratio between the OD of the target sample and the CO). Sample is considered positive when the Index is ≥ 1.0. Horizontal dashed-line represents the limits of detection of IFA (1:20 dilution), vertical line represents the cut-off of positivity for ENZY-WELL ELISAs (Index =1.0). p-value<0.05 was considered statistically significant.

The diagnostic specificity was also calculated by adding to the 115 samples tested by IFA, the 353 samples collected from healthy donors in the pre-pandemic period. Results of the analysis showed an increase of specificity for IgG (96.3%, CI 95%, 94.0-97.8) and IgM (96.9%, CI 95%, 94.5-98.4), while similar data were obtained for IgA (96.3%, CI 95%, 93.9-97.7).

The analysis of the results on 29 samples from COVID-19 patients with known date of symptoms onset (DSO), ranging from 2 to 37 days, showed that the sensitivity of the ENZY-WELL SARS-CoV-2 ELISAs increased after 10 DSO. In detail, 100% sensitivity was reached after 10 and 12 DSO for IgG and IgA, respectively; for IgM, 87.5% was achieved after 12 DSO (**Table 2**).

**Table 2:** Sensitivity according to the days from symptoms onset.

| Days from symptom onset | Sensitivity |  |  |
| --- | --- | --- | --- |
|  | IgG | IgM | IgA |
| <10 | 66.7% | 58.3% | 58.3% |
| >10 | 100.0% | 70.6% | 94.1% |
| >12 | - | 87.5% | 100.0% |

For 40 serum samples (median time of collection: 12 DSO, range: 2-33) parallel nasopharyngeal swab testing had been performed, allowing the evaluation of concomitant presence of SARS-CoV-2 RNA in nasopharyngeal swabs and specific antibodies in serum. The results showed detection of IgA in 70% (n=28), IgG in 75% (n=30) and IgM in 57.5% (n=23) of these samples.

ENZY-WELL SARS-CoV-2 tests showed only a moderate cross-reactivity with serum samples from patients positive for other infectious and non-infectious pathological conditions (**Table 3**). Of note, the majority of cross-reactivity was observed in samples from SARS-CoV cases for all antibody classes, in line with the high similarity between SARS-CoV and SARS-CoV-2.

**Table 3:**
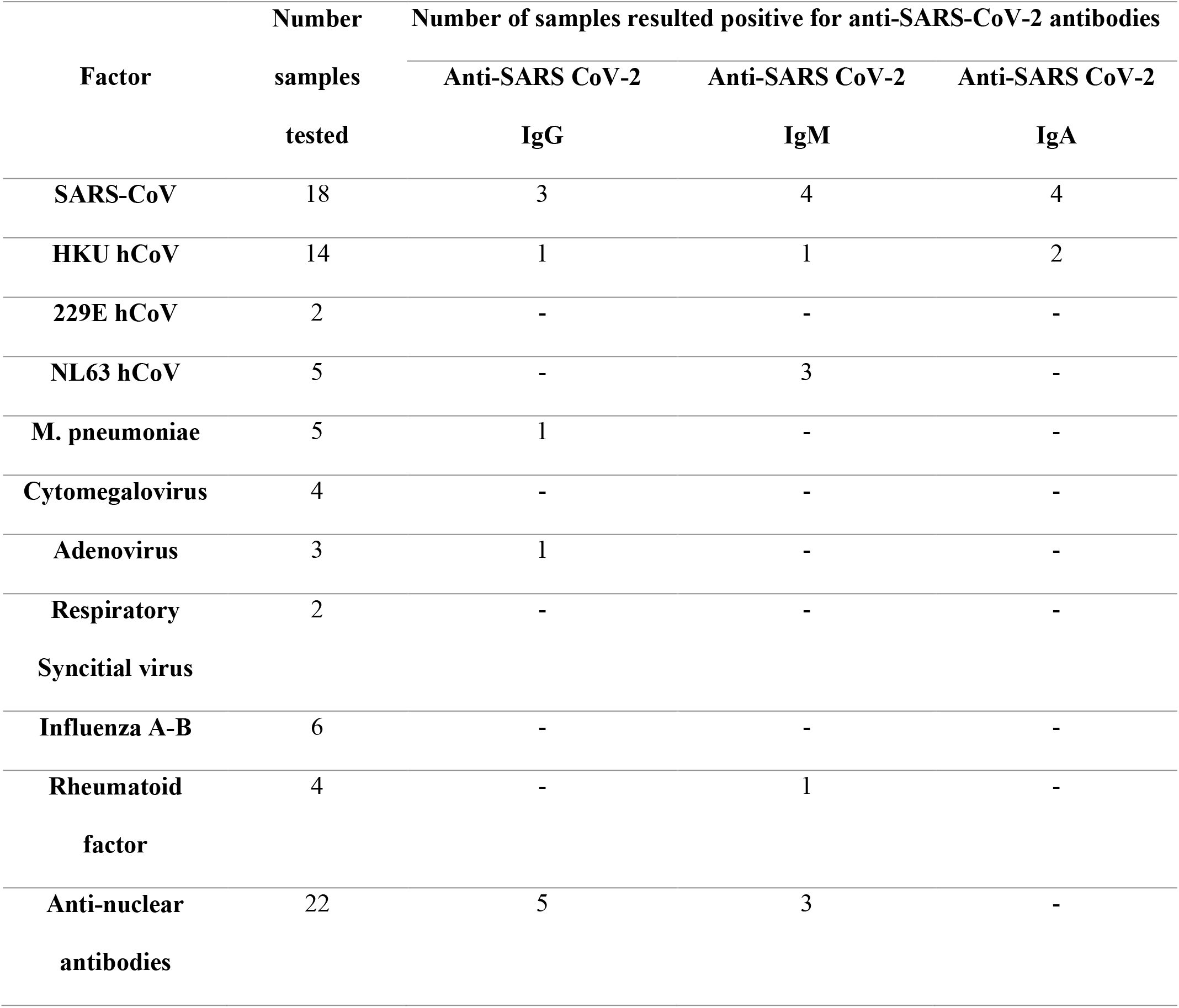
Reactivity with other infectious and non-infectious factors.

## Discusion

In the present study, we report the first validation study of the new commercial whole-virus based ELISAs (ENZY-WELL SARS-CoV-2) detecting anti-SARS-CoV-2 IgM, IgG and IgA.

Overall, diagnostic sensitivity of ENZY-WELL SARS-CoV-2 ELISAs resulted higher for IgG (92.5%) and IgA (93.6%) than IgM (87.7%). Similar results were observed for concordance with IFA, with and excellent agreement for IgG and IgA (0.84 and 0.90, respectively). These results are in line with other reports and support the inclusion of IgA in the panel of antibody testing (**6, 7, 8**), also considering that this antibody class may be pathologically relevant for respiratory infections (**9**). Specificity was similar for all ENZY-WELL SARS-CoV-2 ELISAs (IgG: 96.3%; IgM: 96.9%; and IgA: 96.3%). These tests showed a moderate cross-reactivity to other infectious and auto-immunity factors, which has to be taken into account for the interpretation of results. The main evidence of non-specific reactivity was observed in sera from SARS-CoV cases and for IgM ELISA (**3**). IgA ELISA showed the lowest number of non-specific reactions both to the tested infectious and non-infectious interfering factors.

Differently from other serological tests based on specific viral antigens (such as N or S proteins), these assays are based on crude extract from SARS-CoV-2 whole-virus which can increase the possibility of antibodies detection, especially when the antigenic determinants for anti-SARS-CoV-2 humoral response need to be fully identified. On the other hand, it could affect the specificity, an aspect that need to be evaluated in comparison with single antigen-based tests.

As shown in previous reports, sensitivity is linked to the DSO due to the seroconversion window. In fact, sensitivity of ENZY-WELL SARS-CoV-2 IgG and IgA ELISAs resulted 100% in those samples collected after 10 and 12 DSO, respectively, while 88.5% of sensitivity was obtained for IgM after 12 days (**3, 7**).

In the present study, the ELISAs were performed in manual mode; however, the tests can be totally automatized and carried out from primary samples collection tube in combination with automated ELISA analyzer with Laboratory Information System (LIS). This solution represents a great advantage reducing laboratory turnover, especially when high loads of samples need to be tested, and avoiding operator errors in ELISA procedures.

Laboratory testing plays the major role in identification and monitoring of infected persons, enabling isolation of infected patients and interruption of the chain of transmission. Nucleic acid detection remains the gold standard for identification of acute SARS-CoV-2 infection as the detection of viral RNA in the nasopharyngeal swab or other respiratory specimens is linked to viral shedding and potential for SARS-CoV-2 transmission (**1,4**). Serological testing can support diagnosis and outbreak monitoring, and it can help in understanding immune profiles with implications for epidemiological and public health measures, serologic therapy and vaccine (**1,2**). Therefore, validating new serological methods and determining their performance and potential for cross-reactivity is essential to address the use and ensure proper and reliable diagnosis, clinical and epidemiological. The ENZY-WELL SARS-CoV-2 ELISAs here described may represent a valid option for antibody detection and support its use for both diagnostic and epidemiological purposes, covering all different antibody types developed in SARS-CoV-2 immune response.

## Data Availability

Data available

## Declaration of Competing Interest

Alessandra Brogi, Giulia Tesi and Tommaso Bandini are employees of the DIESSE Diagnostica Senese S.p.a. (Siena, Italy). In no way their contribution to the completion of the study influenced the study design and the analysis of the results. No additional conflict of interest or other competing relationships exist.

## Acknowledgement

This research was carried out in the framework of a scientific collaboration between INMI and DIESSE Diagnostica Senese S.p.a. and supported by Ministry of Health (Ricerca Corrente and Conto Capitale 2017), and European Commission – Horizon 2020 (European Virus Archive GLOBAL - 871029; EU project 101003544 – CoNVat; EU project 101003551 - EXSCALATE4CoV).

We acknowledge the Covid-19 INMI Study Group (Maria Alessandra Abbonizio, Chiara Agrati, Fabrizio Albarello, Gioia Amadei, Alessandra Amendola, Mario Antonini, Raffaella Barbaro, Barbara Bartolini, Martina Benigni, Nazario Bevilacqua, Licia Bordi, Veronica Bordoni, Marta Branca, Paolo Campioni, Maria Rosaria Capobianchi, Cinzia Caporale, Ilaria Caravella, Fabrizio Carletti, Concetta Castilletti, Roberta Chiappini, Carmine Ciaralli, Francesca Colavita, Angela Corpolongo, Massimo Cristofaro, Salvatore Curiale, Alessandra D’Abramo, Cristina Dantimi, Alessia De Angelis, Giada De Angelis, Rachele Di Lorenzo, Federica Di Stefano, Federica Ferraro, Lorena Fiorentini, Andrea Frustaci, Paola Gallí, Gabriele Garotto, Maria Letizia Giancola, Filippo Giansante, Emanuela Giombini, Maria Cristina Greci, Giuseppe Ippolito, Eleonora Lalle, Simone Lanini, Daniele Lapa, Luciana Lepore, Andrea Lucia, Franco Lufrani, Manuela Macchione, Alessandra Marani, Luisa Marchioni, Andrea Mariano, Maria Cristina Marini, Micaela Maritti, Giulia Matusali, Silvia Meschi, Francesco Messina, Chiara Montaldo, Silvia Murachelli, Emanuele Nicastri, Roberto Noto, Claudia Palazzolo, Emanuele Pallini, Virgilio Passeri, Federico Pelliccioni, Antonella Petrecchia, Ada Petrone, Nicola Petrosillo, Elisa Pianura, Maria Pisciotta, Silvia Pittalis, Costanza Proietti, Vincenzo Puro, Gabriele Rinonapoli, Martina Rueca, Alessandra Sacchi, Francesco Sanasi, Carmen Santagata, Silvana Scarcia, Vincenzo Schininà, Paola Scognamiglio, Laura Scorzolini, Giulia Stazi, Francesco Vaia, Francesco Vairo, Maria Beatrice Valli).

We also acknowledge Silvana Verdiani, Veronica Ricci, Simona Toppi, Stefania Tornesi, Marco Vaccaro, Giacomo Sampieri, Claudia Soldatini, Helena Cerutti, Marinunzia Castria, Andrea Ianniello, Francesco Petrini e Alessia Bogi.

## INMI Covid-19 laboratory team

Abbate Isabella, Agrati Chiara, Aleo Loredana, Alonzi Tonino, Amendola Alessandra, Apollonio Claudia, Arduini Nicolina, Bartolini Barbara, Berno Giulia, Biancone Silvia, Biava Mirella, Bibbò Angela, Bordi Licia, Brega Carla, Canali Marco, Cannas Angela, Capobianchi Maria Rosaria, Carletti Fabrizio, Carrara Stefania, Casetti Rita, Castilletti Concetta, Chiappini Roberta, Ciafrone Lucia, Cimini Eleonora, Coen Sabrina, Colavita Francesca, Condello Rossella, Coppola Antonio, D’arezzo Silvia, Di Caro Antonino, Di Filippo Stefania, Di Giuli Chiara, Fabeni Lavinia, Felici Luisa, Ferraioli Valeria, Forbici Federica, Garbuglia Anna Rosa, Giombini Emanuela, Gori Caterina, Graziano Silvia, Gruber Cesare Ernesto Maria, Khouri Daniele, Lalle Eleonora, Lapa Daniele, Leone Barbara, Marsella Patrizia, Massimino Chiara, Matusali Giulia, Mazzarelli Antonio, Meschi Silvia, Messina Francesco, Minosse Claudia, Montaldo Claudia, Neri Stefania, Nisii Carla, Petrivelli Elisabetta, Petroni Fabrizio, Petruccioli Elisa, Pisciotta Marina, Pizzi Daniele, Prota Gianluca, Raparelli Fabrizio, Rozera Gabriella, Rueca Martina, Sabatini Rossella, Sarti Silvia, Sberna Giuseppe, Sciamanna Roberta, Selleri Marina, Selvaggi Carla, Sias Catia, Stellitano Chiara, Toffoletti Antonietta, Truffa Silvia, Turchi Federica, Valli Maria Beatrice, Venditti Carolina, Vescovo Tiziana, Vincenti Donatella, Vulcano Antonella, Zambelli Emma.

